# Routine data collection in home care: A national survey of home care providers in England

**DOI:** 10.1101/2024.06.10.24308698

**Authors:** V Davey, J Healey, J Liddle, B Beresford, S Rand, C Goodman, K Spilsbury, B Hanratty

## Abstract

**Purpose:** Mandatory digital social care records and a standardised schedule for collecting information on home care clients are proposed for regulated adult social care providers in England. This is akin to a minimum dataset (MDS). This study aimed to understand current data collection practices in home care, and identify where support for implementation of an MDS is needed.

**Design:** An online survey of English home care providers was conducted in 2023, asking about the information they collect, store, and share about their clients. Data were analysed using descriptive statistics and logistic regression.

**Findings:** One hundred and fifty-five responses were received from home care providers in all regions of England, a majority were for-profit organisations (89%). All collected a range of data on client characteristics and observations about care delivered. Monitoring of changes in client wellbeing and use of standardised measurement tools (e.g. functioning, mood or quality of life) were uncommon. Over two-thirds (71%) reported that they reviewed the content of care packages at least every six months. Providers with a majority of self-funding clients were more likely to regularly update information on care needs and client/ family preferences.

**Originality:** To the best of our knowledge, this is the first national survey of home care providers on their routine data collection practices.

**Practical implications:** Data collection in UK home care will require expansion, to implement an MDS, which has resource implications for providers. Home care staff will need the skills to collect and use data to enhance client care.

## Introduction

Home or domiciliary care supports almost one million people with long-term care needs in the UK, approximately twice the number of people living in care homes. It has a critical impact on the individuals and families who receive support (Boyle, Seddon, & Toms, 2023; Rand, Forder, & Malley, 2017; Rand et al., 2022) but also accounts for a significant component of public sector spending (Allan et al., 2021; Gridley et al., 2022). Data availability has been a major barrier to increasing our understanding of this important area of care. Home care clients may have physical, mental and/or cognitive impairments, but unlike health service patients, information on their characteristics is not readily available (Curry & Oung, 2021).

The organisation and funding of home care services in England precludes systematic exchange of client information across health and social care services (Author’s own, *In Review*). As a result, organisations have to collate their own information. The content of records at provider-level is loosely specified in the UK, unlike other countries where documentation is often standardised and structured (Mitchell et al., 2023; Morandi et al., 2024; Puustinen, Kangasniemi, & Turjamaa, 2021). At local authority (LA) level there is a standardised approach to collecting data. Each year, local authorities in England use the same survey instrument to sample people receiving social care support and their carers, including people who pay privately for care (known as self-funders). The survey captures a number of outcomes, including the impact of services on the quality of life (QoL) of users and unpaid carers, continuity and quality of care, independence and empowerment (NHSE, 2023a, 2023b).

In the future, ambitious targets mandating digital care records for England are expected to transform the content and availability of data at provider level in social care (DHSC, 2022a, 2022b, 2023). Digital care records systems (DSCRs) will have to comply with international e-health data standards to ensure interoperability among digital systems being used by health providers (NHSE, 2022; NLM, 2023). DSCRs should also be able to integrate with the NHS applications to allow care workers a filtered view of a client’s electronic healthcare record. (NAO, 2018). Data could also be linked at individual-level, using a unique client identifier (e.g. National Health Service or National Insurance number) offering a range of new possibilities for understanding the home care population and their health, care, and support needs over time (Burton et al., 2022). DSCRs are expected to make it possible to obtain data on adult social care directly from providers which would reduce the need for burdensome data collection by local authorities.They may also support person-centred care, with some of the proposed content informed by people with lived experience of care (PRSB 2021).

As greater use is made of client information routinely collected by home care providers, it will be important that data are standardised for analysis and reporting, and suitable for aggregation. In particular, home care providers will need to use standardized measures to record client characteristics and wellbeing. A minimum specification for the client level data collected by home care providers (often termed a Minimum Dataset or ‘MDS’) is expected to be embedded within future DSCRs in England. The potential benefits of populating an MDS with routinely collected data in home care have been recognised (e.g. (Dickins, Joe, & Lowthian, 2023; Morandi et al., 2024), and the need for enhanced data in home care is widely acknowledged. However, despite a plan for rapid implementation of digital records in England, our understanding of current practice is limited. Little is known about what kind of information home care providers currently collect about their clients and how well current practices would support the introduction of an MDS.

## Methods

### Study aims

The survey aims were to describe the data routinely collected by home care providers to establish if the range and content of data currently collected are sufficient to support the move to standardised methods of data capture.

### Study design

We conducted an electronic survey, hosted on the Qualtrics survey platform (Qualtrics, Provo, UT, USA), to ask UK home care providers about the information they collect and store about their clients.

The survey comprised 45 questions, organised into six sections. In this paper, we report on data collected on the following topics: characteristics of respondent home care organisations; client information collected on service entry; frequency of data review; use of satisfaction surveys and/or QoL measures. In a separate article published elsewhere, we describe progress towards, and experiences of, digitalisation amongst home care providers (Healey et al., 2024).

The survey was developed in two stages. A prototype was reviewed by a small number of academics independent of the research team, and representatives of two national organisations representing UK home care providers. A version was then piloted in Qualtrics with senior managers of two home care providers using cognitive interview techniques (Ryan, Gannon-Slater, & Culbertson, 2012). Revisions were made and tested in a second pilot. A copy of the survey questions is provided in Supplementary File #1.

### Data collection

In England there are 11,204 home care providers registered with the regulator Care Quality Commission (CQC), with many providers affiliated to membership bodies. To reach regulated organisations that were providers of regular home visits, an email invitation to take part in the survey (including anonymous hyperlink to the survey and with the study information sheet attached) was distributed by national, regional and local membership bodies representing both for-profit and not-for-profit home care providers. The research team also distributed the email invitation via their existing networks and posted information about the survey on social media. One email reminder was sent out by all routes mentioned above. The online survey was operational between 19th October and 9th December 2022. Survey completion was anonymous, but respondents could volunteer the name of their home care organisation. Instructions requested completion by the owner/director or a manager including obtaining informed consent for participation.

### Data analysis

The data were cleaned in Excel and imported into SPSS (SPSS 25.0) for descriptive analysis and logistic regression to explore relationships between key characteristics of home care organisations and the types of data collected (e.g. funding source, organisation size measured by caseload size, number of operating bases, regional footprint, use of digital records, nature of home care provided).

## Results

### Sample

One hundred and fifty-five responses were received. A majority were from for-profit organisations (n =134, 89%), and independent businesses (n = 94, 60%). Just under one third were care/home care chains (17, 30%), and one fifth “franchise” owners (home care organisations run independently but in accordance with the franchisor branding and standards) (30, 19%) (**Table I**).

**Table I:** Survey of home care providers: Sample characteristics.

|  | Frequency (n) | % |
| --- | --- | --- |
| **Respondent's role within the home care organisation** |  |  |
| Business owner/ Franchisee | 56 (155) | 36 |
| CEO/Executive Director or other Director | 28 (155) | 18 |
| Senior manager | 24 (155) | 15 |
| Registered Manager | 41 (155) | 26 |
| Other | 7 (155) | 4 |
| **Home care provider type** |  |  |
| Franchise* | 30 (155) | 19 |
| Local office/ branch of chain business | 8 (155) | 24 |
| Independent business<sup>†</sup> | 94 (155) | 60 |
| Local authority in-house service | 5 (155) | 5 |
| Chain business (head office) | 9 (155) | 6 |
| Other | 10 (155) | 6 |
| **Scale of home care organisation (by N of branches)** |  |  |
| 1 office/ operating base | 79 (112) | 71 |
| 2-3 branches/ offices | 18 (112) | 16 |
| 4-10 branches/ offices | 8 (112) | 7 |
| 11-19 branches/ offices | 2 (112) | 2 |
| 20+ branches/offices | 5 (112) | 4 |
| **Caseload<sup>§</sup>** |  |  |
| < 20 clients | 21 (152) | 14 |
| 20-49 clients | 37 (152) | 24 |
| 50-99 clients | 34 (152) | 22 |
| 100-249 clients | 29 (152) | 19 |
| 250 and above | 31 (152) | 20 |
| **Spread of home care organisation** |  |  |
| Only one region | 132 (152) | 86 |
| 1-3 regions | 13 (152) | 8 |
| 4 regions and above | 9 (152) | 6 |
| **Financial structure** |  |  |
| For profit | 134 (151) | 89 |
| Not for profit | 16 | 10 |
| Social Enterprise | 0 | 0 |
| Other | 1 | <1 |
| **Services provided** |  |  |
| Regular domiciliary care visits | 145 (155) | 93 |
| Reablement | 51 (155) | 33 |
| Live-in care | 56 (155) | 36 |
| Continuing Health Care (CHC) funded care | 94 (155) | 60 |
| Other | 9 (155) | 6 |
| **Main funding source** |  |  |
| All or mostly LA or NHS funded (incl. DP) | 65 (151) | 43 |
| All or mostly self-funded | 54 (151) | 36 |
| Roughly equal | 32 (151) | 21 |

\*This is a home care agency run independently but in accordance with franchisor branding and standards.
<sup>†</sup>Nationally, 65% of home care organisations registered by CQC are independent businesses (Laing & Buisson 2021).
<sup>§</sup>Nationally 41% of registered home care organisations have < 20 clients, 29% have between 20-49 clients, 17% have been 50-99 clients, 10% have 100-249 clients and 2% have 250 clients and above (Ibid).

Caseloads ranged from fewer than 20, to more than 250 clients. Compared to national regulator data on home care providers, our respondents were more likely to have high or very high caseloads and less likely to be micro-providers (<20 clients). Thirty-eight per cent of our respondents had a caseload below the national average (47 clients), compared to 58% of all providers (LaingBuisson, 2021). From caseload data, we estimate that survey respondents represent around 6.5% of services provided in England. In addition to regular domiciliary care visits, just over a third of respondents were from organisations providing reablement or restorative care (a short-term intervention delivered to people living in their own homes which seeks to restore, or maximise, independence in activities of daily living). A similar proportion were providing 24-hour live-in care. Organisations that offered more than one type of home care were likely to be providing multiple types.

Providers responded from all regions of England, more than half (51%) from London and the Southeast. Responses were in line with variations in regional market fragmentation (LaingBuisson, 2021). Most providers (86%) operated in only one region. The proportion of small providers (up to 49 clients) amongst respondents varied by region, from 42% in London to 14 % in the Northeast. Large providers (> 250 clients) accounted for around one third of the sample in *all* regions, except the Northeast where 50% of the sample had >250 clients.

Organisational size was associated with funding source. Very small (<19 clients) small (20-49 clients) and medium (50-99 clients) organisations were more likely to have a high proportion of clients who had local or national government funding (Local authority or National Health Service (NHS)). Providers were split between those that used all or predominantly digital records (n = 77, 50%) and those that used a mix of digital and paper records (n = 71, 46%). A minority were entirely paper-based (n = 7, 4.5%).

### Data collected at outset and frequency of care package review

Administrative data routinely collected at entry point to a service almost always included the origin of the referral (n = 138, 88%) and funding source (n = 142, 90%). Fewer than half of responding home care providers recorded NHS number and only six in ten recorded a local authority reference number. This practice was associated with funding status, and much more likely to be recorded by home care providers who were mostly reliant on local authority and/ or NHS funding (*X*^*2*^ (1, *N* = 150) = 26.838, p = <0.001). Similarly, organisations that routinely record NHS number were likely to record information on primary care and community health care (*X*^*2*^ (1, *N* = 155) = 5.740, p = 0.017) but were not more likely to record hospital services involved (**Table II**). Organisations that *did not* report recording NHS number, were more likely to only provide standard home (domiciliary) care (*X*^2^ (1, *N* = 155) = 6.142, *p* = 0.013) and also record their clients’ National Insurance number.

**Table II:** Information on health, functioning and care delivery, routinely collected in home care.

| Types of client information collected | Frequency (n) | % |  |  | Frequency of Updating |  |  |
| --- | --- | --- | --- | --- | --- | --- | --- |
|  |  |  | When changes occur (%) | Only at care package review (%) | No standard practice (%) | Not updated | Not stated<sup>1</sup> (%) |
| Information about health conditions and disabilities |  |  |  |  |  |  |  |
| Diagnosed physical health conditions | 148 (155) | 95 | 90 | 3 | - | - | 7 |
| Diagnosed mental health conditions | 144 (155) | 92 | 86 | 3 | - | - | 11 |
| Memory or cognitive impairment | 146 (155) | 94 | 85 | 3 | - | - | 12 |
| Sight or hearing impairment | 143 (155) | 92 | 83 | 4 | - | - | 13 |
| Communication needs | 145 (155) | 93 | 84 | 3 | <1 | - | 12 |
| Diagnosed learning disabilities | 116 (155) | 74 | 65 | 3 | 2 | - | 30 |
| Autism diagnosis | 94 (155) | 60 | 54 | 3 | <1 | - | 43 |
| Information on the involvement of other services and informal carers |  |  |  |  |  |  |  |
| Other social care services | 138 (155) | 89 | 89 | - | - | - | 11 |
| Primary care/ community health care | 132 (155) | 85 | 85 | - | - | - | 15 |
| Health clinics/ services involved | 117 (155) | 75 | 75 | - | - | - | 25 |
| Informal carers | 126 (155) | 81 | 81 | - | - | - | 19 |
| Information about care needs, goals and preferences and care package details |  |  |  |  |  |  |  |
| Care needs (e.g., person care, domestic, mobility) | 140 (155) | 90 | 86 | 3 | 1 | - | 10 |
| Regular medications | 139 (155) | 90 | 88 | 1 | 1 | - | 10 |
| Client/ family desired outcomes/ goals for care | 138 (155) | 89 | 74 | 10 | 4 | <1 | 11 |
| Preferences for how care is delivered | 139 (155) | 90 | 81 | 8 | 1 | - | 10 |
| Care package details | 137 (155) | 88 | 84 | 4 | <1 | - | 12 |
| Information routinely collected in a client's daily care file/ record (paper or digital) |  |  |  |  |  |  |  |
| Care tasks completed at each visit | 153 (155) | 99 | 86 | 3 | 1 |  | 10 |
| Medication prompting and administered (MAR) | 154 (155) | 99 | 88 | 1 | 1 | - | 10 |
| Any healthcare tasks (e.g., wound care, health status monitoring) | 150 (155) | 97 | 74 | 10 | 4 | 1 | 11 |
| Adverse events or incidents | 153 (155) | 99 | 81 | 8 | 1 | - | 10 |
| Observed changes in client's wellbeing (e.g., pain, confusion, mood) | 142 (155) | 92 | 84 | 4 | <1 | - | 12 |

<sup>1</sup>Not stated' includes organisations that stated that they do record the information but did not state when they update, and organisations that did not indicate that they record the information.

Over two-thirds of home care providers (71%, n = 95) reported that they reviewed clients’ care packages every six months, or more often. Organisations providing continuing health care (n = 94, 60%) were more likely to review clients’ packages at least every 6 months (*X*^2^ (1, *N* = 134) = 4.270, *p* = 0.039), while organisations only providing regular domiciliary care were less likely to do so (*X*^2^ (1, *N* = 134) = 6.270, *p* = 0.012). Frequency of client review was not associated with funding source, caseload size, number of operating bases, or use of digital care records.

### Information to support care delivery

Home care providers reported the collection of a wide range of information to support care planning and delivery (**Table II**). This included data on health conditions, disabilities, involvement of other services and unpaid carers, care needs, goals and preferences, care package details and a range of information recorded in a client’s care record as part of daily observations.

Almost all responding home care providers recorded observations about care at each visit, including tasks undertaken, medication prompts/ administration and adverse events. Documentation of perceived changes in psychosocial wellbeing (mood, loneliness or social networks) was seldom recorded in a standardised format (**Table III**). However, most providers record observed changes in wellbeing in aspects *such as* pain, confusion, mood in a non-standardised way (**Table II**). Updating of the latter was variable: sometimes only at care package reviews. Others provided no data on regularity of updates, suggesting that formal documentation was not the primary means for exchanging information on these aspects of client wellbeing. Otherwise, most information to support care delivery was updated when changes occur, but less so for health care tasks. There was no association between this, and the provision of NHS funded ‘continuing health care’.

**Table III:** Home care data collection: Referral, care package review and use of standardised measurement tools.

| How often client packages are reviewed |  |  |  |
| --- | --- | --- | --- |
|  | Frequency (n) | % |  |
| Annually | 39 (135) | 29 |  |
| Every 6 months, or more frequently | 95 (135) | 71 |  |
| Types of client information collected |  |  |  |
| Who made the referral (e.g. client, family, LA) | 138 (155) | 88 |  |
| Who is funding the care (e.g. self, DP, LA) | 142 (155) | 90 |  |
| NHS number | 71 (155) | 45 |  |
| Local Authority reference number | 93 (155) | 60 |  |
| National Insurance number | 19 (155) | 12 |  |
| Routine use of standard measurement tools |  |  |  |
|  | Frequency (n) | % Responding home care organisations | % Care homes (Hanratty et al 2023) |
| Physical Wellbeing |  |  |  |
| Independence/ functioning (e.g. Barthel Index) | 15 (140) | 11 | 85 |
| Frailty (e.g., Clinical Frailty Score) | 19 (140) | 14 | 28 |
| Physical Activity (e.g., Physical Activity Scale for the Elderly - PASE) | 9 (140) | 6 | 84 |
| Sleep (e.g. Sleep Quality Scale) | 7 (140) | 5 | 92 |
| Skin condition (e.g., Waterlow Scale) | 59 (140) | 42 | 85 |
| Nutrition (e.g., MUST screening scale) | 57 (140) | 41 | 68 |
| Psychosocial Wellbeing |  |  |  |
| Mood (e.g. Geriatric Depression Score) | 16 (140) | 11 | 86 |
| Loneliness (e.g., UCLA Loneliness Scale) | 4 (140) | 3 | - |
| Social Networks (e.g., Lubben Social Networks Scale) | 4 (140) | 3 | - |
| Other |  |  |  |
| Pain (e.g. Abbey Pain Scale) | 27 (140) | 19 | 62 |
| Routine use of quality-of-life tools* |  |  |  |
| Adult Social Care Outcomes Tool (ASCOT) | 20 (155) | 13 | - |
| ICECAP Quality of Life Measure | 5 (155) | 3 | - |
| Euro-QoL EQ-5D (or EQ 3-D) | 1 (155) | <1 | - |
| R-Outcomes | 5 (155) | 3 | - |
| Routine use of satisfaction tools |  |  |  |
| Client satisfaction survey/questionnaire | 139 (155) | 89 | - |

\*Other quality of life tools noted were, Personal Outcomes Scale (Ghent) (n=1), Outcomes Wheel (n=1), UDSET (n=1) and other, non-specified (n=3).

Home care providers serving all or a majority of self-funding clients were significantly more likely to report that they regularly update information on care needs, client/ family desired outcomes, goals for care, preferences for how care is delivered and care package details when changes occur (X^2^ (1, N =150) = 4.334, p = 0.037, X^2^ (1, N =150) = 7.454, p = 0.006, X^2^ (1, N =150) = 4.666, p =0.031, X^2^ (1, N =150) = 4.426, p =0.035, respectively). Overall, use of digital records did not appear to be associated with the types of information collected to support care delivery, or how often data items were updated. Providers who used some paper records were more likely to report that that they routinely update information on client/ family desired outcomes, when changes occur (X^2^ (1, N =155) = 7.265, p =0.007).

Client diagnoses of learning disabilities and autistic spectrum disorder were recorded by 75% and 60% of the 155 respondents. Providers with 20-49 clients were more likely to collect these data (X^2^ (1, N =151) = 4.907, p =0.027). This reflects the size of organisations providing care to people living with these conditions.

### Routine use of standardised measures for assessment or monitoring

Routine measurement of independence/ functioning, physical activity and mood was limited, as was data collection on skin integrity and nutrition (**Table III**). Routine observation of pain using standardised measures was reported by just under a fifth of respondents. Medium sized providers (caseloads of 50-99 clients) were less likely than providers of other sizes to use standardised measures of physical wellbeing (independence/ functioning, frailty, physical activity, and sleep) (X^2^ (1, N = 136) = 4.086, p = 0.043), but as likely as any size of organisation to monitor skin integrity or pain using recognised tools.

There was no relationship between the provision of reablement services and recording of data on frailty, independence/ functioning or physical activity. However, it should be noted that the survey only captured home care providers that were providing reablement services in addition to standard domiciliary care services.

Psychosocial aspects of wellbeing (social networks and loneliness) were recorded by just 3% of the sample. Routine collection of measures of psychosocial wellbeing (comprising either/or, mood, loneliness or social networks) was negatively associated with providing live-in care (X^2^ (1, N = 139) = 3.713, p = 0.054) but more frequent among home care providers with extremely small caseloads (<20 clients).

Quality-of-life measures were not being systematically used by the majority of providers. Of the options offered (**Table III**) the most frequently used was the Social Care Related Quality of Life measure (ScRQoL), ASCOT. We received one free text response, describing use of a measure not listed. ScRQoL was in routine use in 13% (n = 20) of responding organisations, which were more likely to have very low caseloads (<20 clients) (*X*^2^ (1, *N* = 130) = 3.848, *p* = 0.050). There was no evidence of any relationship between use of digital care records and routine use of standardised measures of physical wellbeing, psychosocial wellbeing and quality-of-life.

### Routine measurement of satisfaction with the service

Most providers employed client satisfaction surveys, either a bespoke tool (n = 121, 85%), or one developed elsewhere (n = 17, 12%). Development of an in-house client satisfaction survey was positively associated with provider size, as measured by caseload. Organisations with over 250 clients were particularly likely to have developed their own measures (X^2^ (1, N = 139) = 5.297, p = 0.021).

## Discussion

This study focused on collection of information about clients within UK home care, as critical evidence of readiness for a home care MDS. Providers are collecting a range of data on client characteristics and daily observations about care delivery, but few are routinely recording changes over time in client wellbeing using standardised tools. Any home care MDS that contains accurate and up-to-date information on health, care, and support needs and quality of life would require standardised data collection. Our findings suggest an absence in routine use, of standardised measurement tools in home care, ranging from measures of independence/ functioning, physical activity, and mood to physical and psychosocial aspects of wellbeing and quality of life. This has important implications for the implementation of a home care MDS.

In parallel work, examining information collected about residents in care homes as an indicator of feasibility of implementation of a MDS in care homes, Hanratty et al. (2023) have demonstrated that care homes are familiar with a wide range of standardised measurement tools for tracking physical wellbeing, despite having minimal training, akin to home care workers. The lack of their use in home care may reflect the context. In home care, people are more reliant on primary care to address their health needs, and lone workers may have a focus on discrete tasks rather than all aspects of care. Home care workers are also less likely than care home staff to have regular interactions with nursing and other allied health professionals (Hamblin, Burns, & Goodlad, 2023). The type of information that is routinely recorded in home care settings is indicative of the perceived scope and purpose of home care (Author’s own, *In Review*) influenced by the tendency for tightly prescribed contracts in publicly funded home care (Davies et al., 2022) and, arguably, where care is self-funded, reflective of what people want to buy. Priority in data recording is given to areas that may be reviewed by the regulatory body or funders, such as medication management and adverse events. This suggests a discrepancy between current practice and recommendations. For instance, home care providers are bound to “actively encourage feedback about the quality of care” (Health & Social Care Act, 17(2)(e)) but this has not led many providers to embrace routine measurement of quality of life as an indicator of care quality and responsiveness to fluctuating need, despite its promotion at local and national levels in England.

Data collection in home care is influenced by the funding source. Where home care organisations rely more heavily on private clients (self-funders) there is a personalised approach to data capture, emphasising recording and updating client and/or family orientated goals. Among organisations where we might expect a focus on health related information (i.e. those providing NHS funded care), and clients whose care needs frequent review, we found little evidence of any particular emphasis on monitoring the impact of care or client changes over time. Alongside funding source, the size of home care organisations does seem to be important in influencing the content of recorded data, evidenced for example, by pockets of innovative use of quality-of-life tools among very small providers.

Our findings are specific to England, but there are parallels with observations of information collected in home care in other countries. In Norway, home care documentation systems have a limited focus on long-term care needs beyond clinical information, with limited collection of data on psychosocial needs, despite national recommendations (Veenstra, Skinner, & Sogstad, 2020). Likewise in Finland, research has found that information on daily activities is most consistently completed, based on a narrow view of individual needs, despite guidance on integrating and recording the views of older people on planning and delivery of their care (Puustinen, Kangasniemi, & Turjamaa, 2021: e144). Neither country has wholly standardised or digitised formats for collecting data in home care and research on the experiences of home care workers in processing client information via mobile devices is still very limited (Perez et al., 2022; Vasalampi, 2017).

The influence of the implementation of digital social care records (DSCRs) on data collection has yet to be seen. Our findings suggest that in England, the use of digital care records does not appear to be changing the types of information routinely collected, or moving it towards less task-orientated content. This is despite the expected ease of use and potential for shaping recording and updating of information (CQC, 2024). Indeed, we found that recording client and family desired goals for care and monitoring and updating of psychosocial aspects of wellbeing were more common among the very small home care providers and those who were still partly reliant on paper-based systems.

### Limitations

Home care organisations with high caseloads were overrepresented in the sample and small organisations were somewhat underrepresented. Larger organisations may have been more likely to be part of provider networks that circulated the survey and have the capacity to respond. Financially robust organisations are also more likely to embrace digitalisation and the economies of scale that can be realised for example, in back-office costs (LaingBuisson, 2021). The proportion of medium sized provider respondents matched national figures, which is an important achievement as they tend to be reliant on public funding (Davies et al., 2020). This survey offers the first broad insight into routinely collected data in home care in England, and how comprehensive this is. It does not provide information on the perceptions of people receiving home care services and their families, and questions on how the data are used are reported elsewhere (Healey et al 2024). Inconsistencies and variable quality in routinely collected data have been described in the UK (e.g., Brown et al., 2022) and other countries (Puustinen, Kangasniemi, & Turjamaa, 2021; Tshering et al., 2024; Veenstra, Skinner, & Sogstad, 2020).

## Conclusion

A home care MDS embedded within mandated DSCRs could offer an efficient and complete means of monitoring the impact of home care, representing a sea change in how data are collected in English adult social care. Our work suggests that routine data collection on health, wellbeing and quality of life is currently limited in home care. The introduction of mandatory digital social care records and a home care MDS will require profound adjustments in the types of routinely collected data and work to identify which measures are feasible to include. Home care organisations reliant on public funding are amongst the least prepared to implement an MDS. Extensive support for implementation of DSCRs and an MDS is likely to be required, ranging from extending data collection to staff training and promoting a culture of joint working. Independent home care organisations have no history of information sharing across health and social care, which may limit the ability of mandatory digital records to promote integrated care. As this study shows, many organisations do not record their client’s health (NHS) or local authority identifier that would be required for data sharing. Future work is needed to understand how best to promote, support and possibly incentivise the implementation and maintenance of digital records in home care.

## Data Availability

All data produced in the present study are available upon reasonable request to the authors

